# SARS-CoV-2 IgG seroprevalence in blood donors located in three different federal states, Germany, July 2020 to June 2021 – a follow-up

**DOI:** 10.1101/2022.02.11.22270833

**Authors:** Bastian Fischer, Tanja Vollmer, Cornelius Knabbe

## Abstract

Seroprevalence studies can contribute to better assess the actual incidence of infection. Since long-term data for Germany are lacking, we determined seroprevalence of IgG-antibodies against severe acute respiratory syndrome coronavirus 2 (SARS-CoV-2) in 3,759 regular blood donors in three German federal states between July 2020 and June 2021. The IgG seroprevalence was 5.48% (95% confidence interval (CI): 4.77–6.25) overall, ranging from 5.15% (95% CI: 3.73–6.89) in Lower Saxony to 5.62% (95% CI: 4.57–6.84) in North Rhine Westphalia.

## Introduction

Since up to 33.3% of SARS-CoV-2 infections are expected to be asymptomatic or paucisymptomatic [1], it can be assumed that the number of unreported infections is high. Detection of anti-SARS-CoV-2 antibodies can be used to detect previous infections and thus has a tremendous importance concerning broad-based surveillance of coronavirus disease 2019 (COVID 19). Although an acute infection with severe acute respiratory syndrome coronavirus 2 (SARS-CoV-2) is usually verified by PCR, a recent publication suggests a positive identification of anti-SARS-CoV-2 IgG antibodies as an acceptable approach to confirm infection [2]. Our initial published data revealed a low seroprevalence in German blood donors between March and June 2020 [3]. However, data for the further course of the pandemic are lacking but could lead to a more precise overview of the actual incidence of infection in Germany. In this follow-up study we therefore determined anti-SARS-CoV-2 seroprevalence in German blood donors resident in the federal states of North Rhine-Westphalia, Lower Saxony and Hesse over a one-year period from July 2020 – June 2021.

## Results

### Presence of anti-SARS-CoV-2 antibodies in blood-donors

A total of 16,217 residual plasma samples from 3,759 regular blood donors, donated in the period between July 2020 and June 2021, were screened for the presence of anti-SARS-CoV-2 IgG antibodies using the semiquantitative enzyme-linked immunosorbent assay (ELISA) from Euroimmun (Lübeck, Germany) targeting the viral spike protein. Initial seropositive samples were verified by two additional assays from Abbott (Wiesbaden, Germany) and Euroimmun, targeting the viral spike or nucleocapsid, respectively. This approach allowed additional distinction to be made between naturally infected and vaccinated individuals, since the latter do not form antibodies against the nucleocapsid. Samples were obtained from donors located in the three German federal states North Rhine-Westphalia (n□=□1,672), Lower Saxony (n□=□816) and Hesse (n□=□1,271). Overall, we detected anti-SARS-CoV-2 antibodies formed after natural infection in a total of 206/3,759 blood donors (5.48%; 95% CI: 4.77–6.25) in our cohort throughout the one-year study period (Figure 2A). Likewise, the seroprevalence did not differ statistically between the three federal states (p□=□0.628), but incidence was highest in North Rhine-Westphalia (5.62%; 94/1,672; 95% CI: 4.57–6.84), followed by Hesse (5.51%; 70/1,271; 95% CI: 4.32-6.91) and Lower Saxony (5.15%; 42/816; 95% CI: 3.73–6.89). Figure 2B compares the percentage SARS-CoV-2 positivity rate among blood donors (seroprevalence, black bars) with the data officially reported by the Robert Koch Institute (RKI) (PCR detection, light gray bars) for the three German federal-states of North Rhine-Westphalia, Lower Saxony, and Hesse. As can be seen, the relative incidences determined in our study were in most months between July 2020 and June 2021 far above those that could be calculated based on the officially reported cases. With 3.88% (February 2021) and 2.31% (June 2021), the highest seroprevalences were detected at the end of the second- and third corona wave (Figure 2B, light yellow areas) in Germany, respectively. During the period between February and June 2021, the vaccination rate among blood donors increased steadily, with 18.52% (696/3.759; 95% CI: 17.29-19.79) of individuals being vaccinated by the end of the study (Figure S1, Supplement).

**Table:** Number of anti-SARS-Cov-2 seropositive blood donors, total samples tested and resulting positivity rates per month in comparison with those officially reported by the RKI (based on PCR-testing) [4].

|  |  | Blood donors (North Rhine-Westphalia, Hesse, Lowe Saxony) |  |  |  | RKI |
| --- | --- | --- | --- | --- | --- | --- |
|  | Month | Anti-SARS-CoV 2 antibodies |  |  |  | Officially reported<br>PCR positivity-rates (%) |
|  |  | n (positive) | n (samples tested) | Incidence (%) | 95% CI |  |
| 2020 | July | 11 | 1460 | 0.75 | 0.38-1.34 | 0.02 |
|  | August | 14 | 1939 | 0.72 | 0.40-1.21 | 0.05 |
|  | September | 3 | 1501 | 0.2 | 0.04-0.58 | 0.06 |
|  | October | 4 | 1378 | 0.29 | 0.079-0.74 | 0.31 |
|  | November | 5 | 1597 | 0.31 | 0.10-0.73 | 0.65 |
|  | December | 18 | 1251 | 1.44 | 0.85-2.26 | 0.68 |
| 2021 | January | 23 | 1122 | 2.05 | 1.30-3.06 | 0.51 |
|  | February | 41 | 1058 | 3.88 | 2.80-5.22 | 0.26 |
|  | March | 29 | 1623 | 1.79 | 1.20-2.56 | 0.42 |
|  | April | 18 | 996 | 1.81 | 1.07-2.84 | 0.66 |
|  | May | 17 | 1296 | 1.31 | 0.77-2.09 | 0.36 |
|  | June | 23 | 996 | 2.31 | 1.47-3.44 | 0.06 |
| Overall |  | 206 | 16217 | 1.27 | 1.10-1.45 |  |

All donors underwent a medical examination before donation, reported that they did not have current or recent diseases and had no physically detectable symptoms of infection such as fever or an increased leukocyte count. A second retrospective survey for SARS-CoV-2 related symptoms was not conducted.

## Ethical statement

We used exclusively waste material from routine laboratory diagnostics. Samples were collected in accordance with the German Act on Medical Devices for the collection of human residual material. The study was approved by the ethics committee of the medical faculty of the Ruhr University Bochum (AZ: 2022-884).

## Discussion

After the first COVID-19 case occurred in Wuhan, China in December 2019, SARS-CoV-2 spread worldwide, leading the world health organisation (WHO) to officially declare the infectious disease as a pandemic on March 11, 2020. Patients aged >60 years and those suffering from various medical comorbidities are at increased risk for severe disease progression. In contrast, young and healthy individuals are often asymptomatically or paucisymtomatically infected, resulting in many cases not being detected [5]. To evaluate this, we determined the anti-SARS-CoV-2 seroprevalence in a cohort of healthy German blood donors over a one-year period and compared it with the officially reported case numbers of the Robert Koch Institute, the public health institute in Germany. In an initial study, we detected anti-SARS-CoV-2 antibodies in a total of 0.91% (95% confidence interval (CI): 0.58–1.24) of blood donors screened between March and June 2020 [3]. Since seroprevalence data for German blood donors are lacking for the further course of the pandemic, the aim of the present study was to extend the seroprevalence screening for a prolonged period between July 2020 and June 2021. As false-positive measurements could account for a considerable number in populations with a low seroprevalence, initial seropositive measurements were first verified with one additional assay, both targeting the viral spike protein. Of 32 initial positive samples, no material was available for further analysis. Consequently, these samples were not included in the final evaluation. We revised 4.8 % (47/981) initially (equivocal) seropositive tested results, resulting in 902 samples being seropositive using both assays targeting the viral spike protein. As COVID-19 vaccines administered in Germany are based on the spike antigen alone, individuals do not express antibodies against the nucleocapsid [6]. To distinguish between naturally infected and vaccinated individuals, we therefore finally used an assay to detect antibodies directed against the nucleocapsid. During the observation period from July 2020 to June 2021, we detected antibodies attributable to natural infection in a total of 206/3,759 (5.48%; 95% CI: 4.77–6.25) individuals (Figure 1). Anti-SARS-COV-2 seroprevalence steadily increased from December 2020 within our analyzed blood donor cohort, peaking in February 2021 (positivity rate 3.88%, 95% CI: 2.80-5.22) at the end of the second corona wave in Germany (Fig 2B). The enhanced infection rate is most likely explainable due to seasonal effects. Colder temperatures during the winter period lead people to spend more time indoors, which increases viral transmission [7]. The national holidays that occur in Germany at the end of December also facilitate the gathering of larger groups. When delayed seroconversion after SARS-CoV-2 infection is considered, our data are in line with those officially reported by the RKI based on PCR-testing, showing highest positivity-rates between November (0.65%) and December (0.68%) 2020. After seropositivity subsequently decreased within our study group until May 2021, proof of anti-SARS-CoV-2 antibodies has risen again in June 2021 (Fig 2B). The main reason for this observation is likely to be the appearance of the novel delta variant of the virus. In Europe, this variant was first detected in the United Kingdom in April 2021 and subsequently became the dominant SARS-CoV-2 strain in most European countries [8]. Compared to the Alpha-variant that had predominated until then, the new Delta variant was characterized, among other things, by increased transmissibility of up to 40-80% [9, 10]. Although the differences between the three German federal states considered are not significant, we detected the highest rate of SARS-CoV-2 infected individuals over the entire study period in North Rhine-Westphalia (5.62%; 94/1,672; 95% CI: 4.57–6.84), followed by Hesse (5.51%; 70/1,271; 95% CI: 4.32-6.91) and Lower Saxony (5.15%; 42/816; 95% CI: 3.73–6.89) (Figure S2, Supplement). This observation could be explained by differences in population density, which is much higher in North Rhine-Westphalia (526 inhabitants/km^2^), when compared to Hesse (297 inhabitants/km^2^) and Lower Saxony (167 inhabitants/km^2^) [11]. Interestingly, previous publications already revealed an association between population density and transmission of the SARS-CoV-2 virus [12, 13].

**Figure 1:**
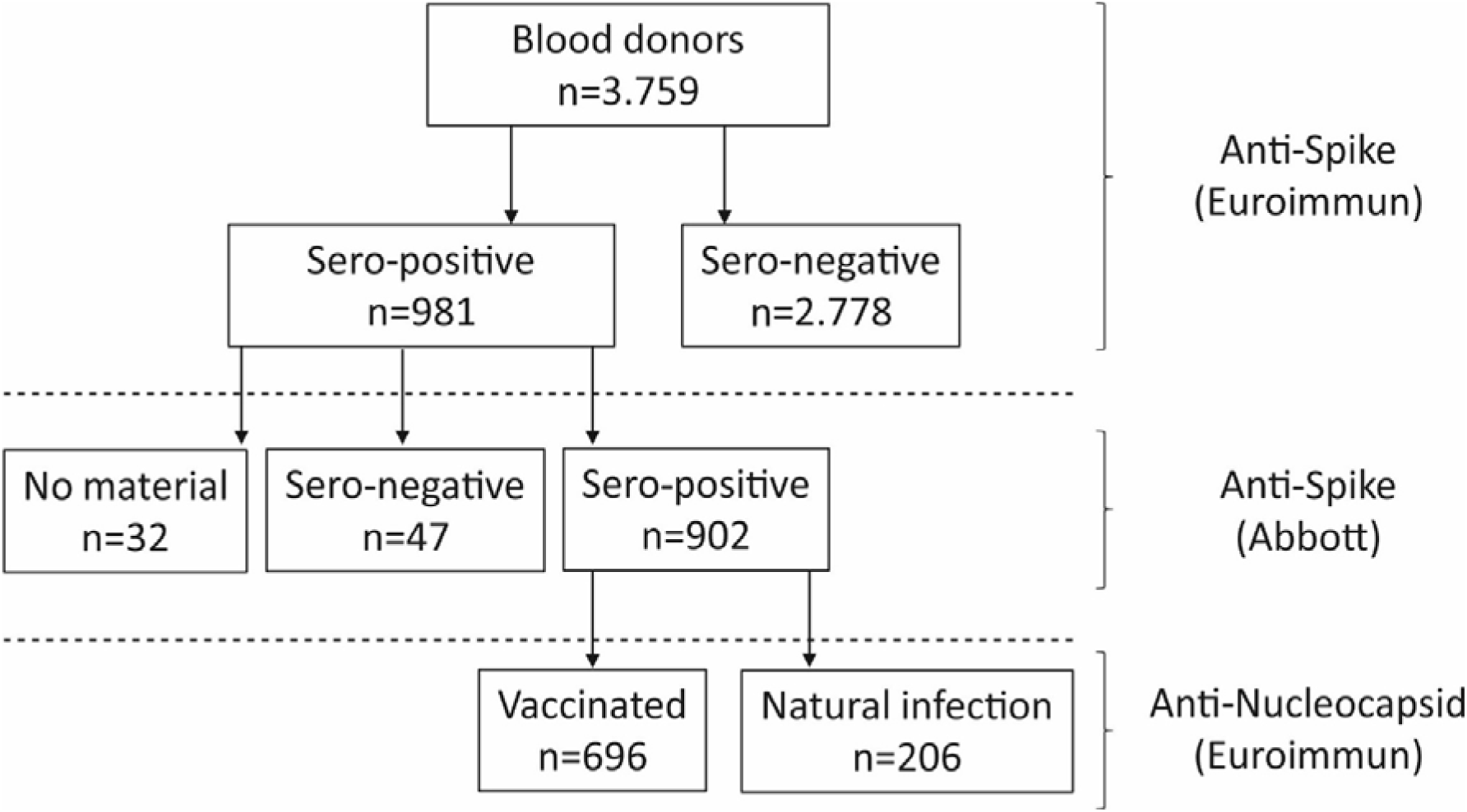
Flow-chart illustrating the methodological design of the present seroprevalence study. A total of 3,759 blood donors were tested for anti-SARS-CoV-2 antibodies over a one-year period. Initial positive samples were validated using a second test. To differentiate between vaccinated and convalescent blood donors among the positive samples, a final assay was performed to detect antibodies against the nucleocapsid, only being expressed by convalescents.

**Figure 2:**
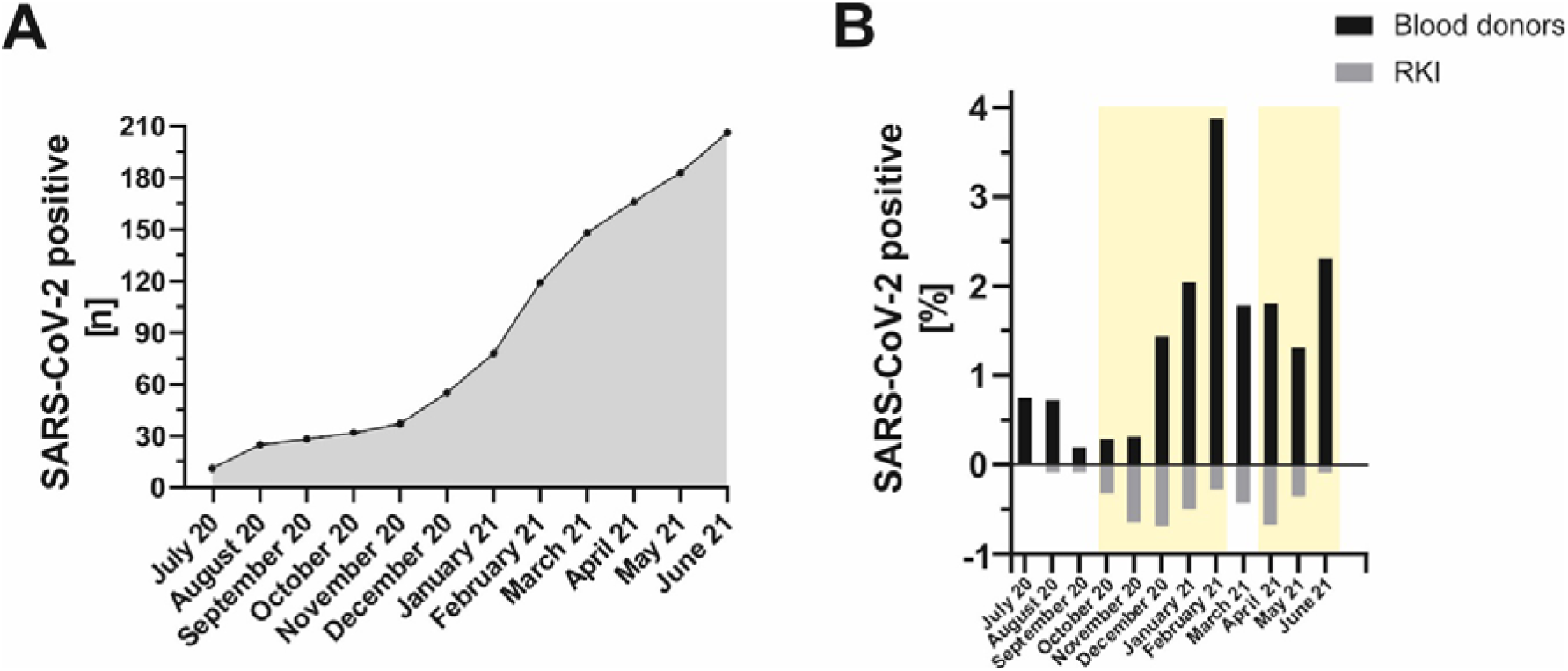
Monthly anti-SARS-CoV-2 antibody detection in the period between July 2020 and June 2021 in blood donors residing in the three German federal states North Rhine-Westphalia, Lower Saxony and Hesse. A: Cumulative illustration of anti-SARS-CoV-2 seropositives by month. B: Comparison of percentage anti-SARS-CoV-2 positivity rate among donors (seroprevalence, black bars) with officially reported data from the Robert-Koch-Institute [4] (PCR detection, grey bars). The light-yellow areas symbolize the second- and third corona wave in Germany.

It should be emphasized that the preselection of blood donors as study cohort is accompanied by limitations regarding representation of the population: Blood donors are between 18 and 65 years-old, young healthy adults are usually overrepresented and other groups (e.g. children, HIV/HCV/HBV-infected patients, older people with underlying conditions, institutionalised people) are excluded or underrepresented. Nevertheless, our findings suggest that there are a large number of unrecorded cases throughout the whole one-year study period. As mentioned in other studies [14], our data therefore reinforces, that longitudinal seroprevalence studies are helpful to better assess the actual incidence of infection within the respectively considered region.

## Conclusion

Our long-term anti-SARS-CoV-2 seroprevalence data of German blood donors help to better evaluate the number of people who have recovered from COVID-19 and elucidate a high number of unrecorded cases.

## Data Availability

All data produced in the present study are available upon reasonable request to the authors.

