## Supplement for "SARS-CoV-2 IgG seroprevalence in blood donors located in three different federal states, Germany, July 2020 to June 2021 – a follow-up"

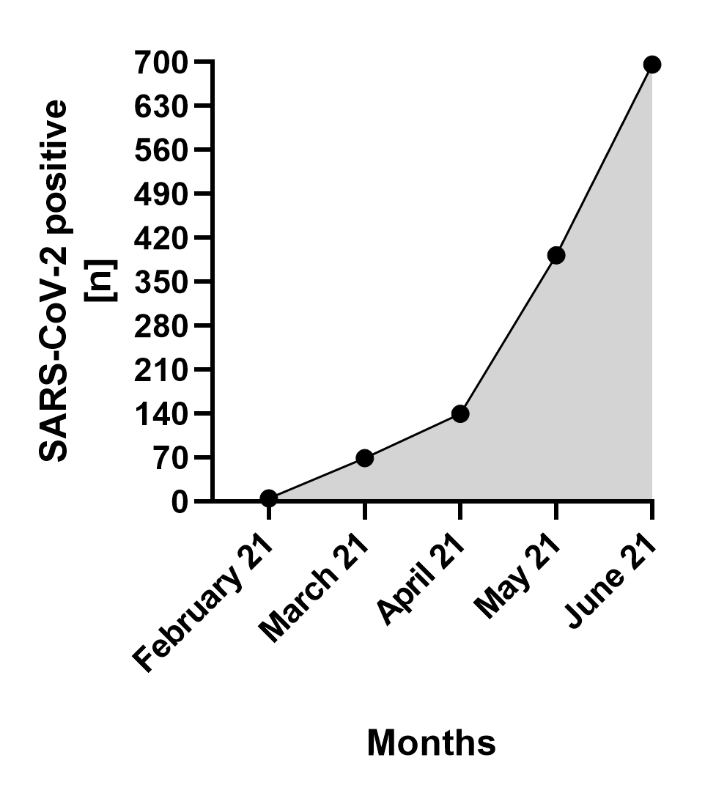

Figure S1: Cumulative illustration of individuals vaccinated against SARS-CoV-2 by month.

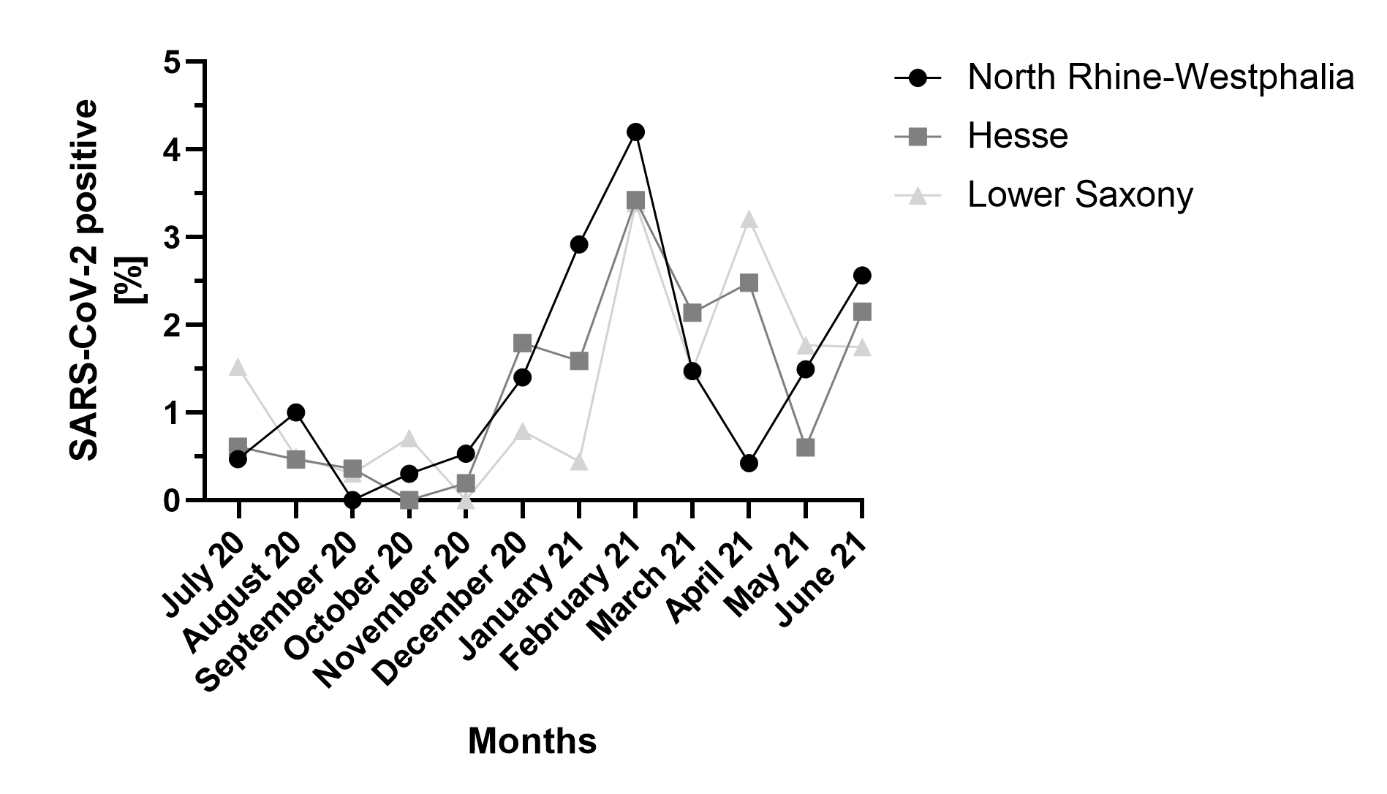

Figure S2: Monthly anti-SARS-CoV-2 antibody detection in the period between July 2020 and June 2021 in blood donors residing in the three German federal states North Rhine-Westphalia (black circles), Hesse (dark-grey squares) and Lower Saxony (light-grey triangles).
